# Impact of speeding on the validity of patient-reported outcome measures. An analysis of psychometric properties stratified by response times

**DOI:** 10.1101/2025.09.24.25335962

**Authors:** Jens Laigaard, Saber Muthanna Saber Aljuboori, Søren Overgaard, Karl Bang Christensen

## Abstract

**Background:** Randomised trials and meta-evidence increasingly rely on patient-reported outcome measures (PROMs). The psychometric properties of patient-reported outcome measures PROMs, including validity, reliability, and responsiveness, are typically established using high-quality datasets, which may not reflect the data quality in clinical trials. Increasing survey burden and fatigue may lead to issues with short response times that can reflect insufficient engagement. This phenomenon, often referred to as ‘speeding’ can lead to random, patterned, or otherwise invalid responses.

**Objective:** This study aims to investigate how response times influence the psychometric properties of the Western Ontario and McMaster Universities Osteoarthritis Index (WOMAC) pain domain in patients with chronic postsurgical pain.

**Methods:** The study is based on responses to the 5-item Western Ontario and McMaster Universities Osteoarthritis Index (WOMAC) pain domain (Likert-scale, version 3.1) from 2,031 patients who underwent total hip arthroplasty (THA), 2,172 patients who underwent total knee arthroplasty (TKA), and 870 patients who underwent unicompartmental knee arthroplasty (UKA) more than one year previously. Each of the three datasets, containing patients who have undergone THA, UKA and TKA, are individually stratified into response time deciles. For each of the deciles, we will evaluate if the data fit a congeneric measurement model, i.e. a model that assumes that the set of observed items all measure the same underlying latent factor. This evaluation of construct validity is done using Item Response Theory (IRT) and Confirmatory Factor Analysis (CFA).

**Perspective:** The results will be submitted for publication in a peer-reviewed journal. We will seek to make the report freely available, either by open-access publication or through publication on a preprint server, e.g. www.medrxiv.org.

## Introduction

Randomised trials and meta-evidence increasingly rely on patient-reported outcome measures (PROMs). PROMs are often criticised for lack of validity. Therefore, validation is needed to increase confidence in trial evidence.

The psychometric properties of patient-reported outcome measures PROMs, including validity, reliability, and responsiveness, are typically established using high-quality datasets.(1) Incomplete and non-content responses (e.g., patterned responses such as straight-lining) may be excluded to ensure that assumptions of the model used for psychometric validations are met.(2) While this approach ensures robust validation, it also introduces a mismatch when PROMs are deployed in clinical research. In practice, all responses, including those potentially compromised by careless or hurried answering, are often retained for analysis. This discrepancy raises concerns about the validity of findings in clinical settings, where response quality may vary widely, and the prevalence of non-content responses is often unknown.

Response time – the duration a participant spends answering each item – has emerged as a potential indicator of response quality, although it is also affected by factors such as age and experience.(3) Short response times may reflect insufficient engagement, leading to random, patterned, or otherwise invalid responses that threaten the integrity of PROM data.(4) However, the impact of rushed responses on the measurement properties of PROMs in a clinical dataset remains poorly understood. A common clinical setting where clinicians rely on PROMs, is the assessment of pain intensity in patients with chronic postsurgical pain after joint arthroplasty.

We therefore aim to investigate how response times influence the psychometric properties of the Western Ontario and McMaster Universities Osteoarthritis Index (WOMAC) pain domain in patients with chronic postsurgical pain.

## Methods

### Data source

In 2023, the author group undertook two surveys of measures of pain and satisfaction 12-18 months after primary THA, TKA or UKA. The surveys were registered at ClinicalTrials.gov (identifiers NCT05845177 and NCT05900791) and listed at the Capitol Region of Denmark’s research (identifiers P-2022-933 and P-2023-4). The primary study populations were patients operated for primary osteoarthritis, namely 1148 UKA patients (870 respondents), 3086 TKA patients (2172 respondents), and 2777 THA patients (2031 respondents). However, a smaller number of patients operated for non-primary osteoarthritis were also surveyed. The median time from surgery to response was 17 months (interquartile range (IQR) 16-18 months) for THA patients, and 13 months (IQR 12-14 months) for UKA and TKA patients. Among other outcomes, the surveys included the the 5-item Western Ontario and McMaster Universities Osteoarthritis Index (WOMAC) pain domain (Likert-scale, version 3.1).(5) *What amount of knee pain have you experienced the last week during the following activities?* a) *Walking on a flat surface, b) Going up or down stairs, c) At night while in bed, d) Sitting or lying, e) Standing upright*. Response options: None; Mild; Moderate; Severe; Extreme. Each of the five items is scored from 0 to 4. These scores are added to a 0-20 sum score, with a higher score indicating worse pain.

### Data imputation

For this study, we will use the full WOMAC pain domain responses. We will apply multiple imputation to impute missing item responses, however empty rows will be omitted from the dataset. We will use chained equations to generate the imputation-enriched dataset using predictive mean matching with the mice package in R (R Core Team, 2023).(6) The imputation model will include both demographic (baseline) and other outcome variables.

### Stratification by response times

Each of the three datasets, containing patients who have undergone THA, UKA and TKA, are individually stratified into response time deciles. That is, the 10% fastest patients who underwent THA are grouped with the 10% fastest UKA and TKA patients, resulting in 10 groups with equal proportions of each surgery type, ordered by response times.

### Reliability

First, we will evaluate if there is **flooring or ceiling effect**, i.e. if many respondents report the lowest or highest possible sum score, respectively. The flooring or ceiling effect is considered substantial if more than 15% of respondents have reported the lowest/highest possible score. Second, we will inspect histograms to evaluate if the distribution of sum scores. Finally, we will assess if the Cronbach’s alpha suggests high internal reliability (i.e. α ≥ 0.70). We chose to also assess Cronbach’s alpha because it is a widely used and interpretable index of reliability for reflective scales. However, with the more detailed evaluation of construct validity using confirmatory factor analysis (CFA), Cronbach’s alpha may be considered superfluous.

### Confirmatory factor analysis (CFA)

Next, we evaluate if the data fit a congeneric measurement model, i.e. a model that assumes that the set of observed items all measure the same underlying latent factor. This is done using **item response theory (IRT**) and **CFA** testing

1. if the instrument is **unidimensional**, i.e. that all items of the instrument measures the same latent variable. This is evaluated using correlations, visualised in a correlation matrix (heatmap), and CFA based on the
  - **Chi**^**2**^**-test**, which tests if the model-implied covariance matrix is significantly different from the observed covariance matrix. Should be insignificant (p>0.05), although biased against large datasets.
  - **Comparative Fit Index (CFI)** (i.e. how well does the factor model fit, compared to a baseline model assuming no relationships among the items). Should be CFI≥0.95.
  - **Root Mean Square Error of Approximation (RMSEA**) which essentially measures how different the proposed factor model is from ‘perfect fit’ to the data. Should be RMSEA≤0.06
  - **Standardized Root Mean Square Residual (SRMR**) which measures how well the model-implied correlations match the actual (observed) correlations among the items. Should be SRMR≤0.08.
2. if there is a **monotonous relationship between the individual items and the latent variable**, i.e. the sum score. Each item is plotted against the sum of the remaining items (**rest-score plots)**. As the score of an individual item increases, the sum score should also increase, with no reversals or inconsistencies. Furthermore, estimated factor loadings from CFA are inspected.
3. if there is no **differential item functioning (DIF)**, i.e. at the same sum score, each items functions similarly across age/sex/etc. We will use **Ordinal Logistic Regression DIF**, to evaluate uniform and non-uniform DIF for each item. Uniform DIF means, that one group scores better/worse across all levels of the latent trait. Non-uniform DIF means that the direction or magnitude of the item bias changes at different levels of the latent trait.

1) if there is **local independence**, i.e. once we control for the sum score, responses to different items should be statistically independent of each other. This is tested similarly to DIF, using Yen’s Q3 residual correlations, plotted to a Residual Correlation Matrix. Q3 residuals > 0.20 above the average often indicate local dependence.

## Tables and figures

Table 1: A table with each response time decile groups demographics, item missingness, and WOMAC pain domain sum score.

Table 2: Construct validity indices, including Cronbach’s alpha and CFA fit indices (Chi^2^-test, CFI, RMSEA, SRMR, a note describing whether there is a monotonous relationship between the individual items and the latent variable, and a note on whether if there is local independence between items), stratified by culture (i.e. data source). For the notes, we will reference to the rest-score plots and residual correlation matrices where the information derives from (supplementary appendices). Mock table:

**Figure 1.**
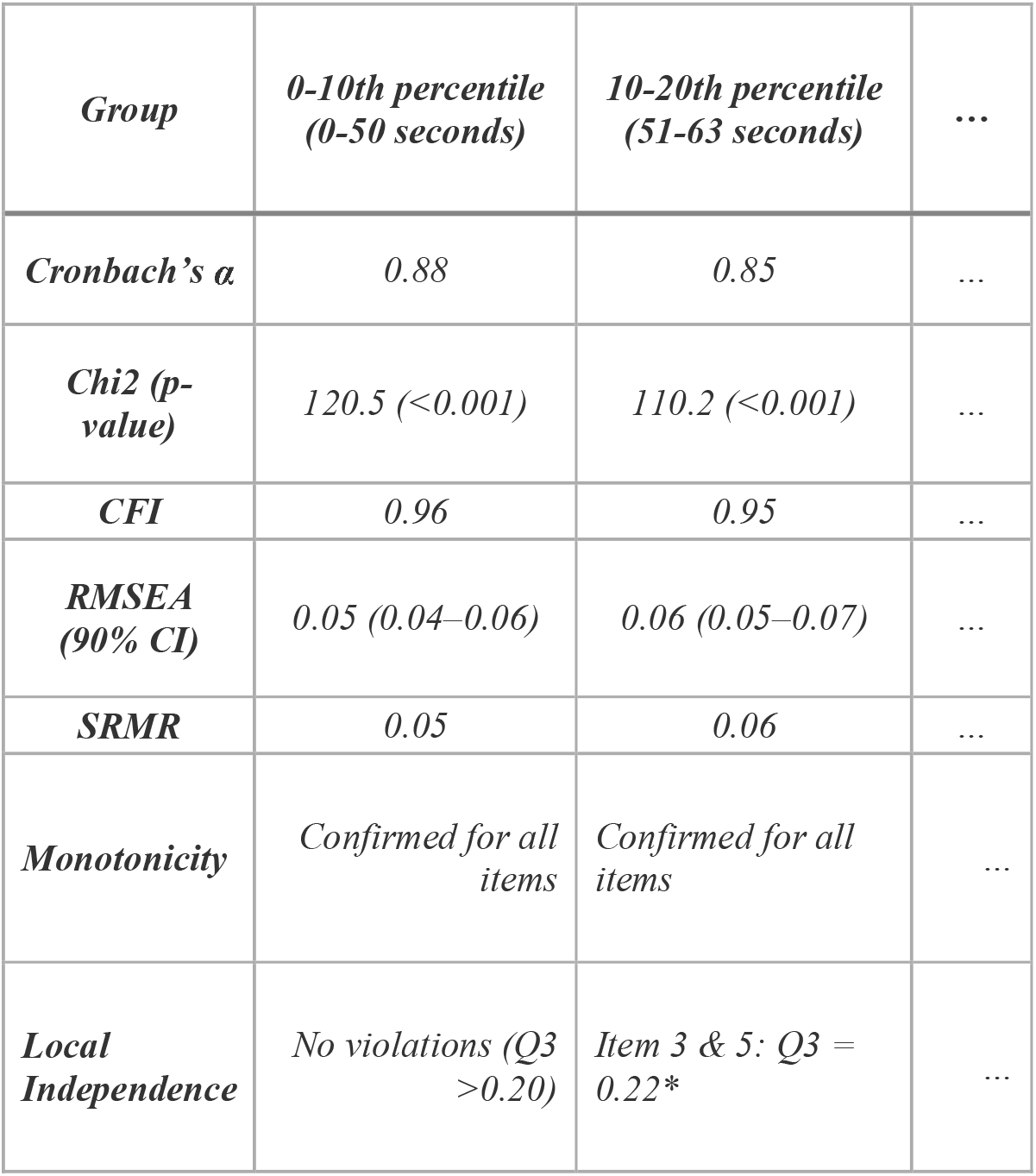
Flow of participant response data.

## Knowledge dissemination

The results will be submitted for publication in peer-reviewed journals. We will seek to make the report freely available, either by open-access publication or through publication on a preprint server, e.g. www.medrxiv.org.

## Ethical Considerations

The project is listed on the Capitol Region of Denmark’s research listing (p-2025-19417), which by delegation from The Danish Data Protection Agency approves the handling of confidential data for research. According to Danish legislation, approval from the national ethics committee is not required for studies that do not collect biological samples or impose interventions (Appendix 1)

Only Jens Laigaard will have access to the data, which are stored, handled and analysed pseudonymised at a logged and encrypted drive. For studies using data from the Danish National Prescription Registry, data are uploaded to Statistics Denmark’s secure online research platform in order to link the data.(7) This remote-access environment can only be accessed with the corresponding author’s unique digital signature.

## Data availability statement

The data used in this study are available from their primary sources upon reasonable request. Please see the original publications for details.

## Appendix 1 – Exempt for notification

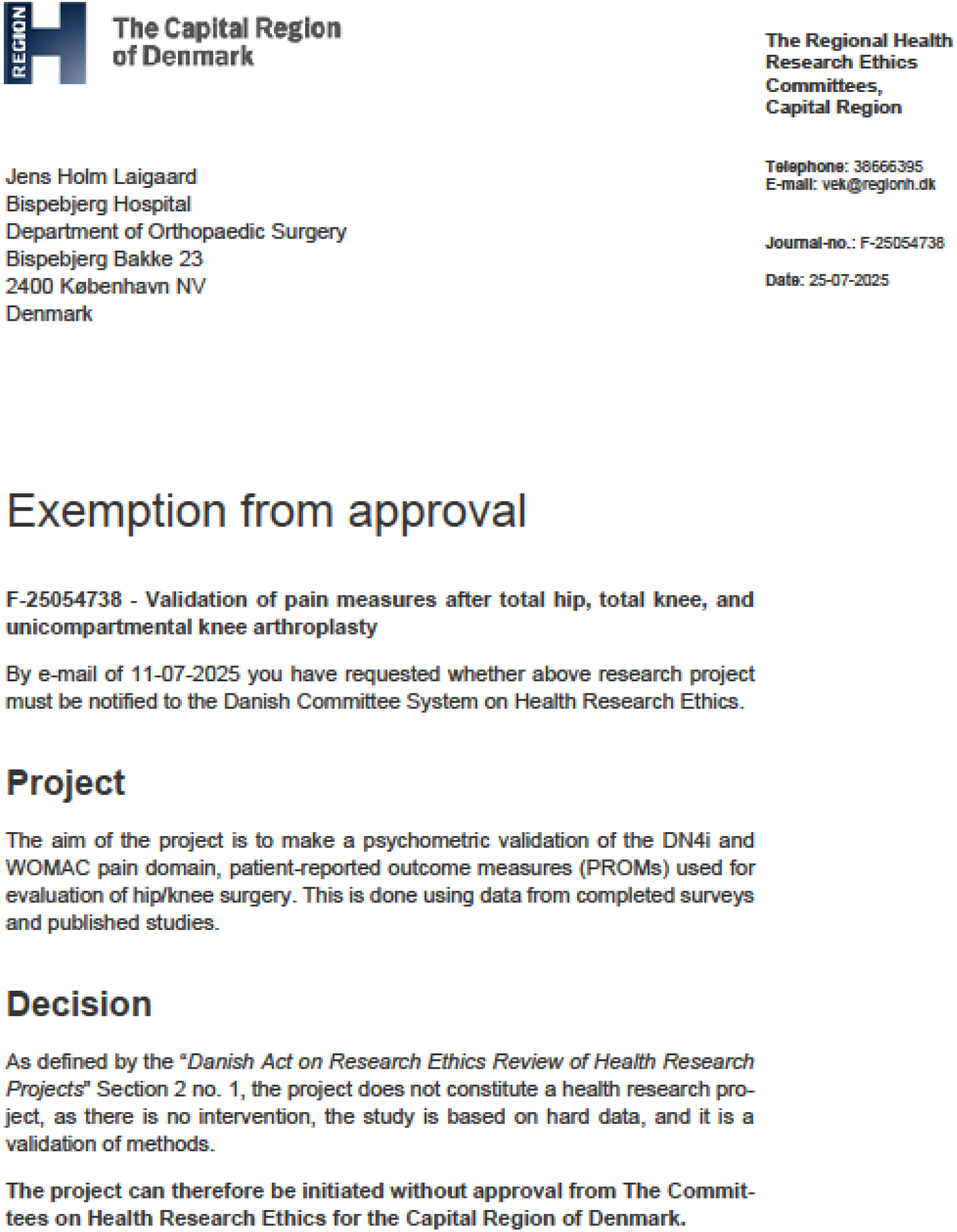

## References

1. Mokkink LB, de Vet HCW, Prinsen CAC, Patrick DL, Alonso J, Bouter LM, et al. COSMIN Risk of Bias checklist for systematic reviews of Patient-Reported Outcome Measures. Qual Life Res. 2018;27(5):1171–9.

2. Gandløse JS, Christensen SWM, Lambertsen DF, Árnason ÓE, Vela J, Palsson TS. Validity and reliability of the Danish version of the Short Form Brief Pain Inventory. Musculoskeletal Science and Practice. 2025 Feb 1;75:103242.

3. Yan T, Tourangeau R. Fast times and easy questions: the effects of age, experience and question complexity on web survey response times. Applied Cognitive Psychology. 2008;22(1):51–68.

4. Zhang C, Conrad F. Speeding in Web Surveys: The tendency to answer very fast and its association with straightlining. Survey Research Methods. 2014 Jul 23;8(2):127–35.

5. Bellamy N, Buchanan WW, Goldsmith CH, Campbell J, Stitt LW. Validation study of WOMAC: a health status instrument for measuring clinically important patient relevant outcomes to antirheumatic drug therapy in patients with osteoarthritis of the hip or knee. J Rheumatol. 1988 Dec;15(12):1833–40.

6. Buuren S van, Groothuis-Oudshoorn K, Vink G, Schouten R, Robitzsch A, Rockenschaub P, et al. mice: Multivariate Imputation by Chained Equations [Internet]. 2025 [cited 2025 Aug 14]. Available from: https://cran.r-project.org/web/packages/mice/index.html

7. Pottegard A, Schmidt SAJ, Wallach-Kildemoes H, Sorensen HT, Hallas J, Schmidt M. Data Resource Profile: The Danish National Prescription Registry. Int J Epidemiol. 2017 Jun 1;46(3):798–798f.

